# Divergent uric acid responses to a traditional Japanese diet and the DPP-4 inhibitor alogliptin in drug-naïve subjects with type 2 diabetes

**DOI:** 10.64898/2026.02.21.26346799

**Authors:** Eiji Kutoh, Alexandra N. Kuto, Nozomi Urushibara, Rumiko Okada, Sachiko Ito

**Author notes:** Address for correspondence: Eiji Kutoh, M.D., Ph.D., Biomedical Center, 1-5-8-613 Komatsugawa, Edogawa-ku 132-0034, Tokyo, Japan.

## Abstract

Uric acid (UA) is traditionally regarded as a metabolic risk marker; however, its dynamic behavior during glucose-lowering therapy remains incompletely understood. We compared UA responses to a modified traditional Japanese diet (MJDD) and the DPP-4 inhibitor alogliptin in patients with early-stage type 2 diabetes mellitus (T2DM). In this prospective observational study, drug-naïve patients received MJDD (n=58) or alogliptin (n=52) monotherapy for 3 months. Changes (Δ) in serum UA were analyzed in relation to glycemic control, insulin resistance, adipose tissue insulin resistance (adipo-IR), and beta-cell function. Both interventions significantly reduced fasting blood glucose and HbA1c while paradoxically increasing serum UA and HOMA-B. Baseline UA was the primary determinant of ΔUA in both cohorts. MJDD significantly reduced body mass index, insulin, free fatty acids, HOMA-R, and adipo-IR, with effects most pronounced in subjects with baseline BMI >25. In contrast, alogliptin selectively reduced adipo-IR in leaner subjects (BMI <25). Across both treatments, ΔUA correlated positively with ΔHOMA-B and inversely with ΔHbA1c. Notably, during MJDD, ΔUA showed a paradoxical negative correlation with ΔBMI and ΔFBG, and a positive correlation with ΔFFA. Patients exhibiting the greatest UA increases demonstrated the most marked improvements in beta-cell function and, with MJDD, the greatest weight loss. These findings indicate that MJDD and alogliptin exert distinct metabolic effects in early T2DM, yet both link rising UA to enhanced beta-cell function, suggesting that UA may serve as a dynamic pharmacometabolic biomarker reflecting therapy-specific metabolic adaptation rather than metabolic deterioration.

## Introduction

Uric acid (UA), the end-product of purine metabolism, has been viewed as a byproduct of impaired renal clearance and oxidative stress through mechanisms such as NADPH oxidase activation, mitochondrial dysfunction, endothelial impairment, and RAAS stimulation (1, 2). Its association with T2DM and metabolic syndrome is well established (3, 4). However, the impact of therapeutic interventions on UA dynamics remains poorly characterized, despite growing recognition that UA is not merely a passive biomarker but a metabolically active molecule influencing cardiovascular and renal outcomes. Weight loss generally lowers UA levels, presumably via decreased production and enhanced renal excretion (5).

The traditional Japanese diet, rich in vegetables, fish, low fat and moderate carbohydrates, is known to improve overweight, glucose control, and insulin resistance (6). Yet, its effect on UA regulation has not been systematically explored. Conversely, DPP-4 inhibitors, widely used in T2DM management, are weight-neutral and do not affect insulin resistance. However, some agents in this class have been associated with UA elevation and improved beta-cell function (7, 8). Interestingly, one of the drugs in this class, alogliptin may exert unique metabolic benefits beyond glycemic control, including reductions in atherogenic lipids and adipose tissue insulin resistance (9), raising the question of whether these effects extend to UA metabolism.

UA has traditionally been linked to insulin resistance and obesity, with elevated levels observed in early T2DM. While improved insulin sensitivity is often assumed to reduce UA through enhanced renal clearance^1^, emerging data suggest a more complex, context-dependent regulation influenced by therapy type and baseline metabolic phenotype (10).

This study compared the effects of modified traditional Japanese diet for diabetes MJDD (11) and the DPP-4 inhibitor alogliptin on UA regulation over three months in subjects with treatment naïve subjects with T2DM, examining relationships with glycemic control, (adipose tissue) insulin resistance, and beta-cell function. We also assessed whether baseline weight modulates these effects. We hypothesized that UA responses reflect distinct, therapy-specific pathways beyond weight reduction or insulin sensitivity alone. These insights may improve our understanding of metabolic heterogeneity and inform individualized treatment strategies.

## Subjects and Methods

### Subjects

The study was conducted in accordance with the Declaration of Helsinki and was approved by the institutional review board (IRB) of Gyoda General Hospital and Kumagaya Surgery Hospital. Written informed consent was obtained and recorded electronically. This project is an extension of the registered study (ID: UMIN000006860), which is listed in the UMIN database. Newly diagnosed, treatment naive subjects with T2DM were enrolled according to Japan Diabetes Society criteria, as described previously (7, 8). Inclusion criteria: HbA1c 6.5–12%, no emergency conditions (e.g., ketoacidosis). Exclusion criteria: renal impairment (creatinine ≥1.5 mg/dL), hepatic dysfunction (AST/ALT ≥70 IU/L), heart disease, severe hypertension (>160/100 mmHg), T1DM, or pregnancy. The subjects were recruited from Gyoda General Hospital, Kumagaya Surgery Hospital (Saitama, Japan), and other affiliated sites of the first author (EK) between December 2015 and January 2025. The drop-outs were excluded from analysis. The final cohort included 110 subjects, assigned to either a modified Japanese diabetic diet (MJDD, n=58) or DPP-4 inhibitor alogliptin (n=52). No other interventions were used. Alogliptin was given at 12.5 mg/day for women and 25 mg/day for men. The subjects were not strictly randomized; this is a comparison of two observational cohorts.

### Modified Japanese Diabetic Diet (MJDD)

The details of MJDD have been published^11^. Briefly, this diet provided 25–30 kcal/kg/day, emphasized fresh vegetables, fruits, daily fish or seafood, limited red/processed meats (≤3 times/week), avoided added oils, encouraged fermented foods, minimized dairy, followed a set meal order (vegetables → protein → carbohydrates), promoted green tea rather than coffee, and slow eating. All participants were encouraged to exercise regularly. The alogliptin group also followed standard dietary advice.

#### 2.3. Laboratory Measurements

The primary endpoint was change in UA after 3 months. The secondary endpoints included FBG, HbA1c, BMI, insulin, FFA, HOMA-R, adipo-IR, and HOMA-B. Fasting blood was collected before breakfast. HbA1c and FBG were measured monthly; insulin at baseline and 3 months (Abbott Japan). Anti-GAD antibodies were checked to exclude T1DM (Mitsubishi BML).

Calculations [11 Kutoh et al., references therein]:

HOMA-R = (insulin × FBG) / 405

Adipo-IR = FFA × Insulin

HOMA-B = (insulin × 360) / (FBG – 63)

#### 2.4. Data Analysis

Changes were calculated as baseline-to-3-month differences. Paired t-tests assessed intra-group changes; simple regression evaluated correlations. Multiple regression identified predictors of ΔUA, with ΔUA as dependent and baseline UA, BMI, FBG, HbA1c, insulin, FFA, HOMA-R, adipo-IR, HOMA-B, T-C, log(TG) and HDL-C as independent variables. The results are presented as mean±SD. Significance was set at p<0.05; p value between 0.05 and 0.1 was considered suggestive. Analyses were performed using PAST software (https://www.nhm.uio.no/english/research/resources/past/index.html) from University of Oslo

## Results

### Baseline Characteristics and Changes in Diabetic Parameters After 3-Month Treatment with the modified Traditional Japanese Diet for Diabetes (MJDD) and DPP-4 Inhibitor Alogliptin

Table 1 summarizes the baseline characteristics and changes in diabetic parameters for each treatment (1A: MJDD; 1B: alogliptin). The baseline levels of diabetic parameters were comparable between these two groups, with no statistically significant differences (data not shown in Table). Both interventions led to significant reductions in FBG, HbA1c or adipo-IR and increases in serum UA and HOMA-B. Reductions in BMI, insulin, FFA or HOMA-R were observed only with MJDD.

**Table 1.**
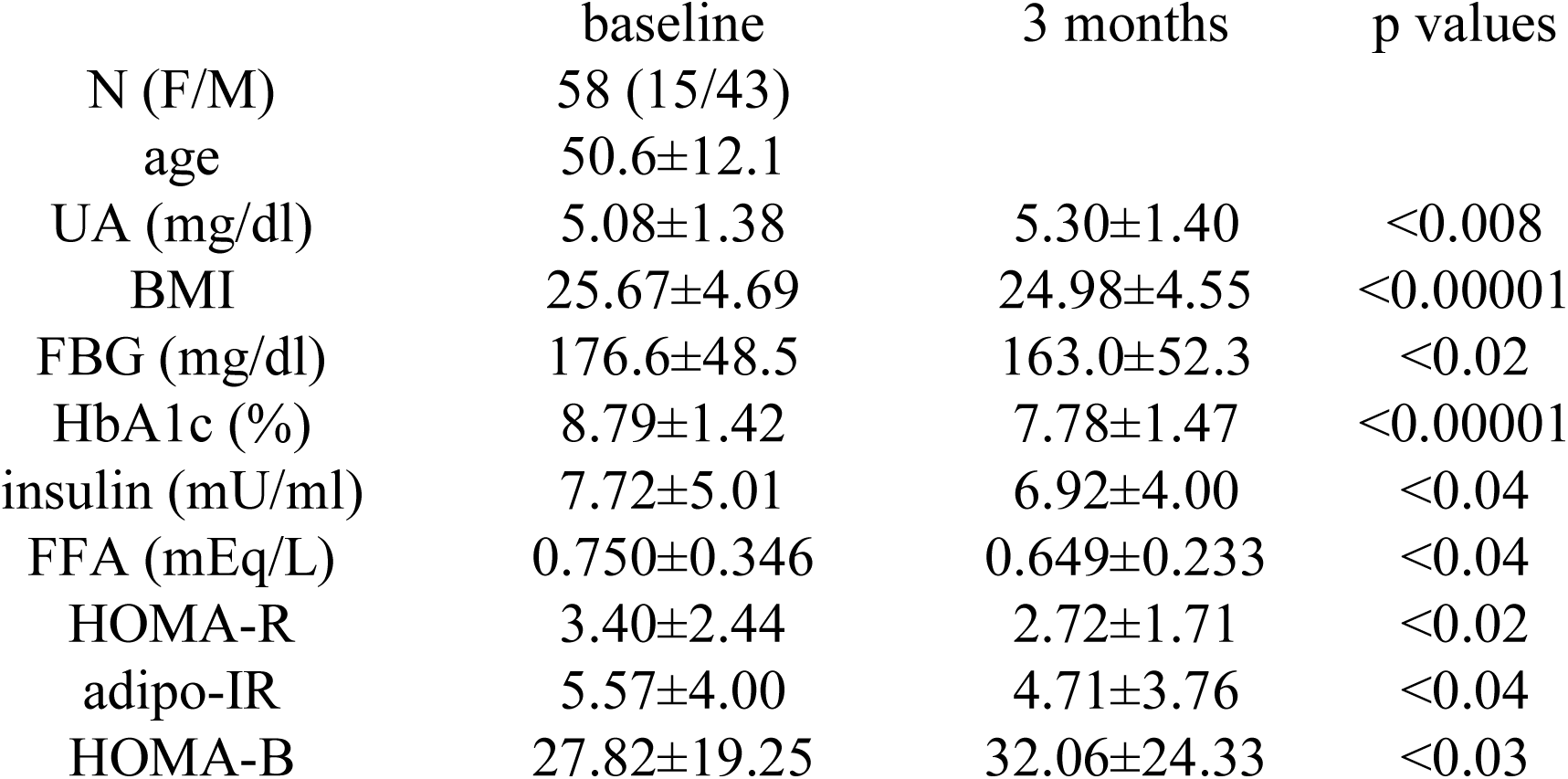
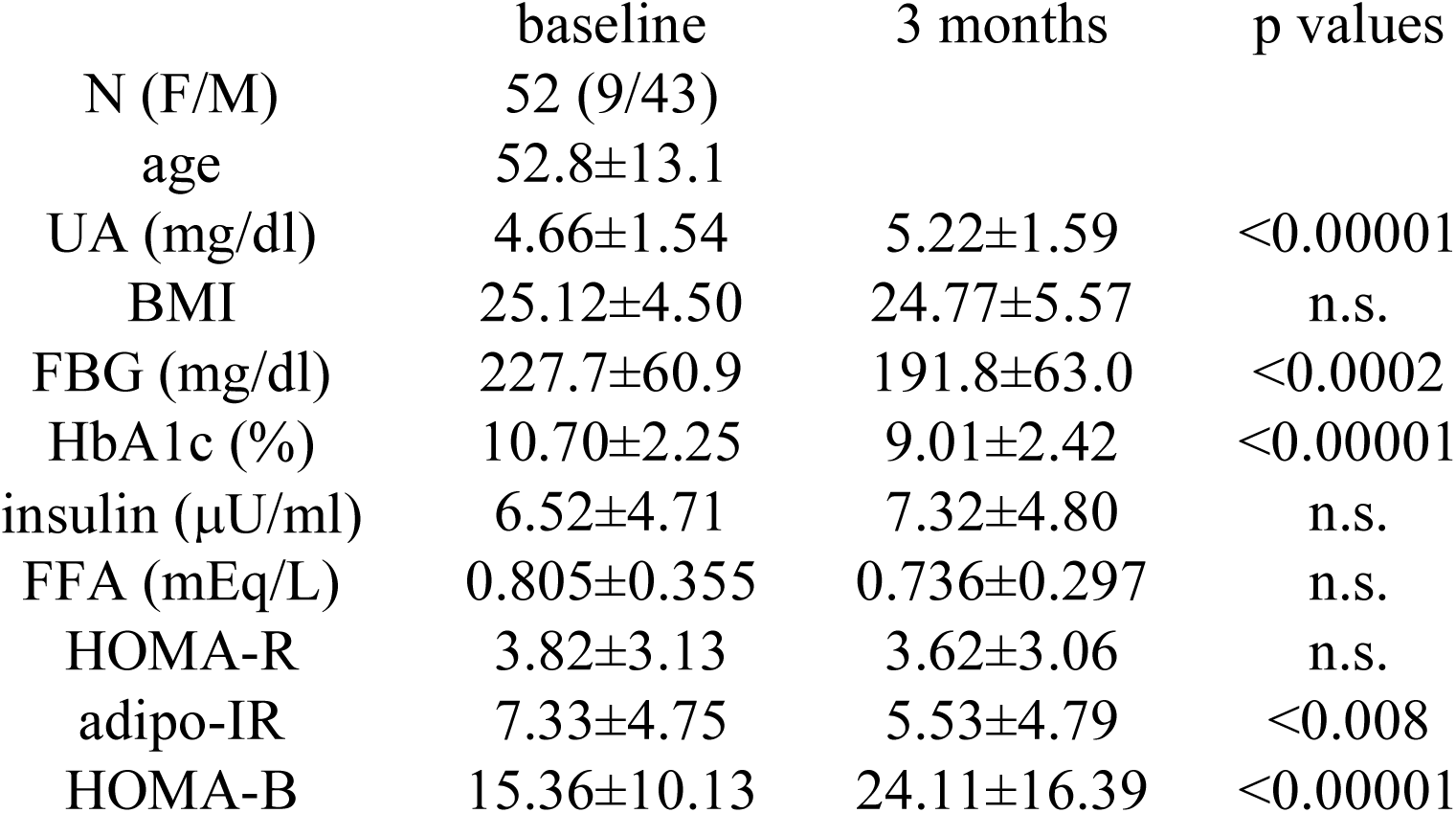
Changes of diabetic parameters with MJDD (modified traditional Japanese diet for diabetes) or DPP-4 inhibitor alogliptin. Paired Student’s t-test was used to compare the changes in the indicated parameters at baseline and 3 months treatment with MJDD or alogliptin. The results are expressed as the mean ± SD (standard deviation). A) MJDD B) alogliptin

|  | baseline | 3 months | p values |
| --- | --- | --- | --- |
| N (F/M) | 58 (15/43) |  |  |
| age | 50.6±12.1 |  |  |
| UA (mg/dl) | 5.08±1.38 | 5.30±1.40 | <0.008 |
| BMI | 25.67±4.69 | 24.98±4.55 | <0.00001 |
| FBG (mg/dl) | 176.6±48.5 | 163.0±52.3 | <0.02 |
| HbA1c (%) | 8.79±1.42 | 7.78±1.47 | <0.00001 |
| insulin (mU/ml) | 7.72±5.01 | 6.92±4.00 | <0.04 |
| FFA (mEq/L) | 0.750±0.346 | 0.649±0.233 | <0.04 |
| HOMA-R | 3.40±2.44 | 2.72±1.71 | <0.02 |
| adipo-IR | 5.57±4.00 | 4.71±3.76 | <0.04 |
| HOMA-B | 27.82±19.25 | 32.06±24.33 | <0.03 |
| N (F/M) | 52 (9/43) |  |  |
| age | 52.8±13.1 |  |  |
| UA (mg/dl) | 4.66±1.54 | 5.22±1.59 | <0.00001 |
| BMI | 25.12±4.50 | 24.77±5.57 | n.s. |
| FBG (mg/dl) | 227.7±60.9 | 191.8±63.0 | <0.0002 |
| HbA1c (%) | 10.70±2.25 | 9.01±2.42 | <0.00001 |
| insulin (μU/ml) | 6.52±4.71 | 7.32±4.80 | n.s. |
| FFA (mEq/L) | 0.805±0.355 | 0.736±0.297 | n.s. |
| HOMA-R | 3.82±3.13 | 3.62±3.06 | n.s. |
| adipo-IR | 7.33±4.75 | 5.53±4.79 | <0.008 |
| HOMA-B | 15.36±10.13 | 24.11±16.39 | <0.00001 |

### Identification of Factors Associated with Changes in UA

Multiple regression analysis was performed to identify baseline factors associated with changes in (Δ)UA, using ΔUA as the dependent variable and baseline UA, BMI, FBG, HbA1c, insulin, FFA, HOMA-R, HOMA-B, T-C, log(TG) and HDL-C as independent variables. With both strategies, baseline UA was selected as a potential predictor for the changes of UA (Table 2A: MJDD, 2B: alogliptin).

**Table 2.**
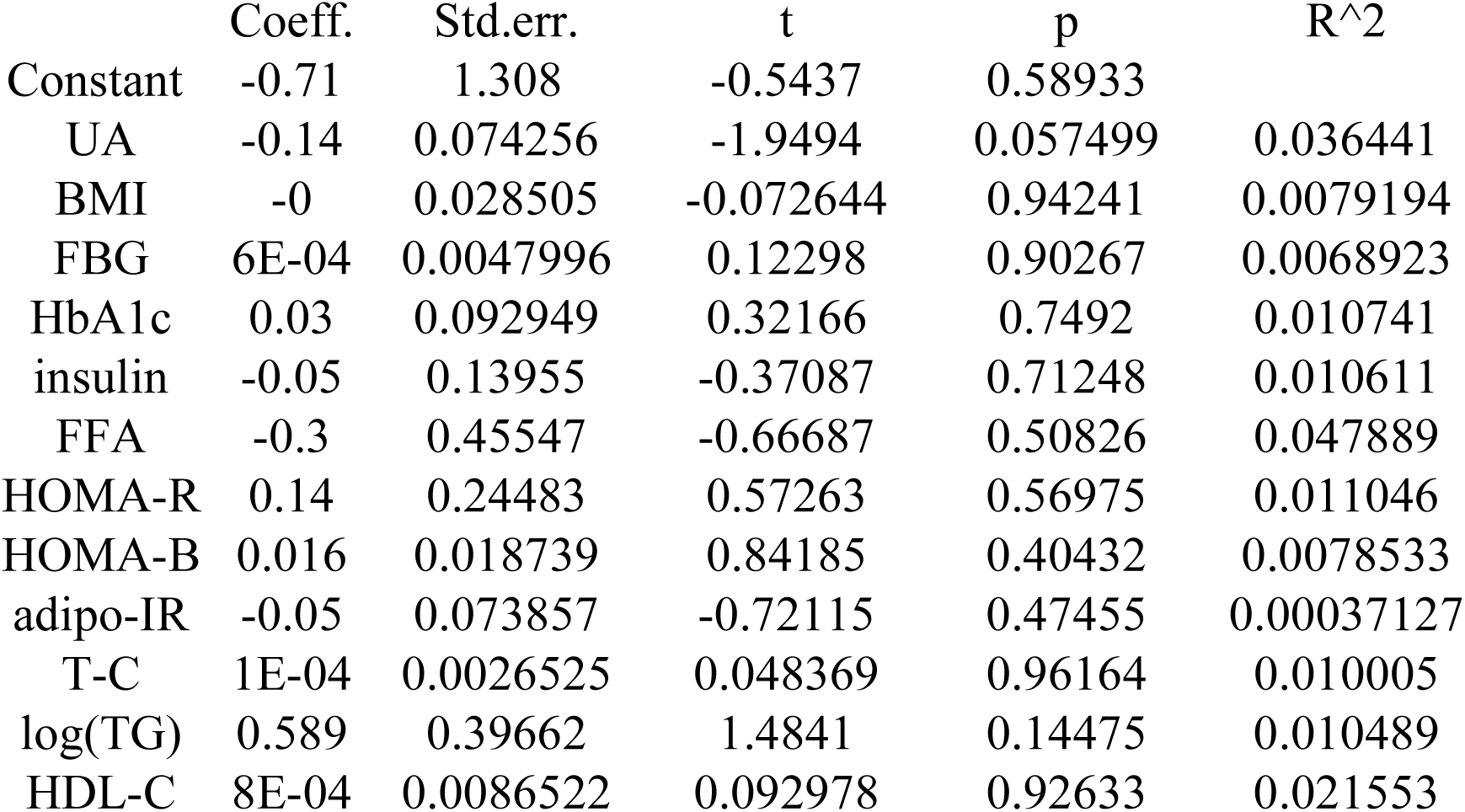
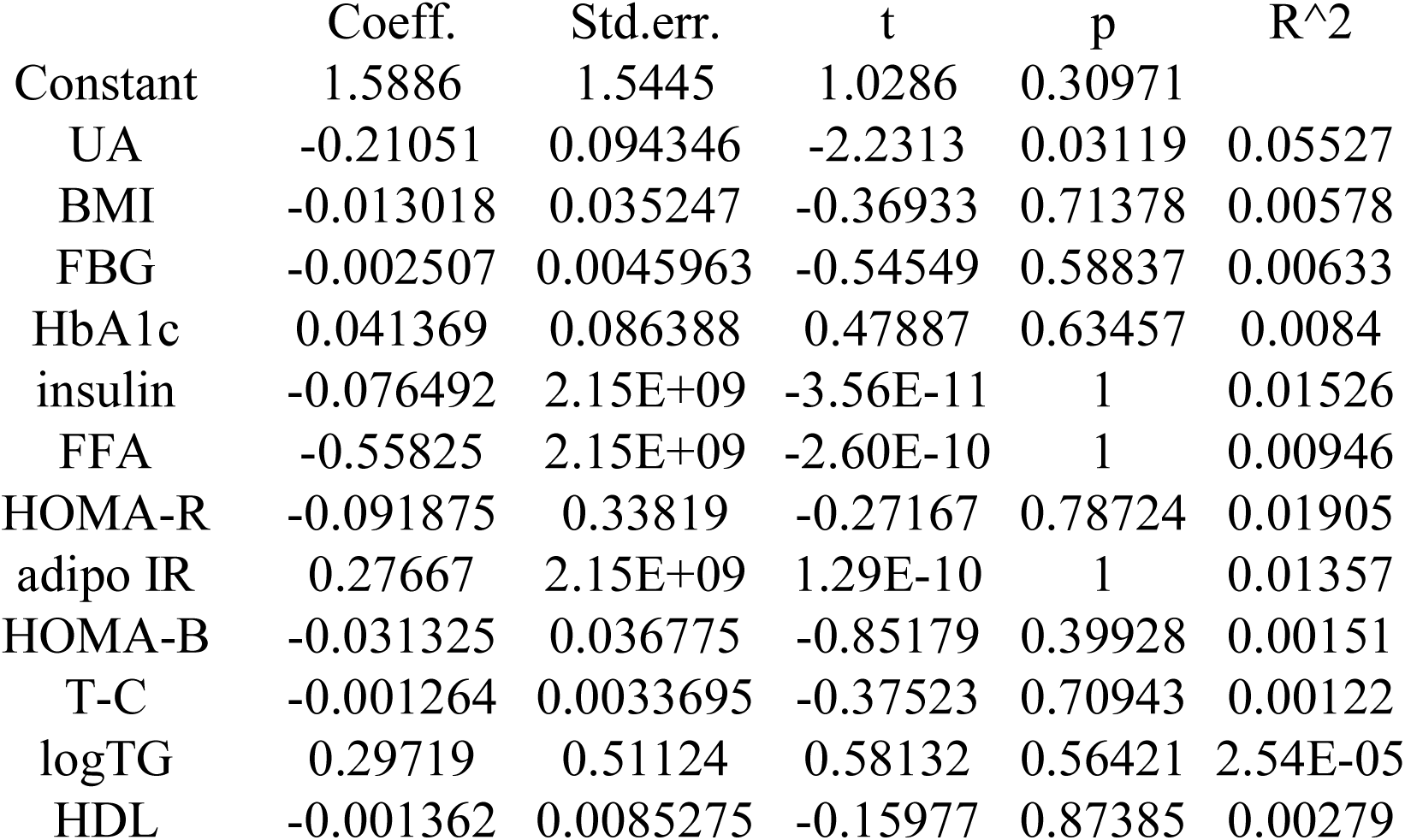
Identification of factors associated with the changes of UA during MJDD and DPP-4 inhibitor alogliptin. Multiple regression analysis was performed using the changes of (Δ) UA as a dependent variable and other parameters including baseline, UA, BMI, FBG. HbA1c, insulin, FFA, HOMA-R, HOMA-B, adipo-IR, T-C, log(TG), and HDL-C as independent variables. A) MJDD B) alogliptin

|  | Coeff. | Std.err. | t | p | R^2 |
| --- | --- | --- | --- | --- | --- |
| Constant | -0.71 | 1.308 | -0.5437 | 0.58933 |  |
| UA | -0.14 | 0.074256 | -1.9494 | 0.057499 | 0.036441 |
| BMI | -0 | 0.028505 | -0.072644 | 0.94241 | 0.0079194 |
| FBG | 6E-04 | 0.0047996 | 0.12298 | 0.90267 | 0.0068923 |
| HbA1c | 0.03 | 0.092949 | 0.32166 | 0.7492 | 0.010741 |
| insulin | -0.05 | 0.13955 | -0.37087 | 0.71248 | 0.010611 |
| FFA | -0.3 | 0.45547 | -0.66687 | 0.50826 | 0.047889 |
| HOMA-R | 0.14 | 0.24483 | 0.57263 | 0.56975 | 0.011046 |
| HOMA-B | 0.016 | 0.018739 | 0.84185 | 0.40432 | 0.0078533 |
| adipo-IR | -0.05 | 0.073857 | -0.72115 | 0.47455 | 0.00037127 |
| T-C | 1E-04 | 0.0026525 | 0.048369 | 0.96164 | 0.010005 |
| log(TG) | 0.589 | 0.39662 | 1.4841 | 0.14475 | 0.010489 |
| HDL-C | 8E-04 | 0.0086522 | 0.092978 | 0.92633 | 0.021553 |
| Constant | 1.5886 | 1.5445 | 1.0286 | 0.30971 |  |
| UA | -0.21051 | 0.094346 | -2.2313 | 0.03119 | 0.05527 |
| BMI | -0.013018 | 0.035247 | -0.36933 | 0.71378 | 0.00578 |
| FBG | -0.002507 | 0.0045963 | -0.54549 | 0.58837 | 0.00633 |
| HbA1c | 0.041369 | 0.086388 | 0.47887 | 0.63457 | 0.0084 |
| insulin | -0.076492 | 2.15E+09 | -3.56E-11 | 1 | 0.01526 |
| FFA | -0.55825 | 2.15E+09 | -2.60E-10 | 1 | 0.00946 |
| HOMA-R | -0.091875 | 0.33819 | -0.27167 | 0.78724 | 0.01905 |
| adipo IR | 0.27667 | 2.15E+09 | 1.29E-10 | 1 | 0.01357 |
| HOMA-B | -0.031325 | 0.036775 | -0.85179 | 0.39928 | 0.00151 |
| T-C | -0.001264 | 0.0033695 | -0.37523 | 0.70943 | 0.00122 |
| logTG | 0.29719 | 0.51124 | 0.58132 | 0.56421 | 2.54E-05 |
| HDL | -0.001362 | 0.0085275 | -0.15977 | 0.87385 | 0.00279 |

### Regulation of Diabetic Parameters According to Baseline BMI

As stated in Introduction, body weight is closely associated with UA levels. The subjects were divided into two subgroups based on (BMI<25, group X, n=29) or (BMI≥25, group Y, n=29). With MJDD, in group X (Table 3A), significant decreases in BMI or HbA1c and increases in UA and HOMA-B were observed. Insignificant decreases of FFA were seen. In group Y (Table 3A), significant decreases in BMI, FBG, HbA1c, insulin, adipo-IR and HOMA-R were seen. A significant inter-group difference in the magnitude of reductions of BMI and HbA1c were observed (greater reduction in group Y vs. group X, Figure 1A, 1B).

**Table 3.** Regulations of diabetic parameters depending on the baseline weight (BMI) with MJDD. The subjects were divided into two groups based on BMI (<25: group X and ≥25: group Y) in these two therapeutic groups. Paired Student’s t-test was used to compare the changes in the indicated parameters at baseline and 3 months treatment. The results are expressed as the mean±standard deviation (SD) A) MJDD B) alogliptin

BMI<25 : group X
|  | baseline | 3 months | p values |
| --- | --- | --- | --- |
| N | 29 |  |  |
| age | 55.5±10.5 |  |  |
| UA (mg/dl) | 4.68±1.34 | 4.92±1.30 | <0.04 |
| BMI | 22.18±1.72 | 21.71±1.79 | <0.03 |
| FBG (mg/dl) | 168.1±45.9 | 159.3±52.7 | n.s. |
| HbA1c (%) | 8.54±1.65 | 7.64±1.38 | <0.0004 |
| insulin (μU/ml) | 5.31±3.43 | 5.35±2.70 | n.s. |
| FFA (mEq/L) | 0.705±0.256 | 0.621±0.260 | 0.096 |
| HOMA-R | 2.19±1.52 | 2.06±1.10 | n.s. |
| adipo-IR | 3.87±3.25 | 3.33±2.52 | n.s. |
| HOMA-B | 21.48±14.96 | 25.18±15.44 | <0.05 |

BMI≥25: group Y
|  | baseline | 3 months | p values |
| --- | --- | --- | --- |
| N | 29 |  |  |
| age | 45.6±11.9 |  |  |
| UA (mg/dl) | 5.47±1.34 | 5.67±1.42 | n.s. |
| BMI | 29.17±4.07 | 28.22±4.17 | <0.00001 |
| FBG (mg/dl) | 185.2±50.4 | 166.6±52.6 | <0.005 |
| HbA1c (%) | 9.03±1.11 | 7.92±1.57 | <0.00001 |
| insulin (μU/ml) | 10.13±5.23 | 8.48±4.50 | <0.009 |
| FFA (mEq/L) | 0.796±0.447 | 0.678±0.204 | n.s. |
| HOMA-R | 4.60±2.62 | 3.38±1.96 | <0.0002 |
| adipo-IR | 7.27±4.01 | 6.09±4.30 | <0.05 |
| HOMA-B | 34.16±21.16 | 38.94±28.93 | n.s. |

BMI<25 : group X
|  | baseline | 3 months | p values |
| --- | --- | --- | --- |
| N | 26 |  |  |
| age | 56.6±13.0 |  |  |
| UA (mg/dl) | 3.98±1.25 | 4.60±1.33 | <0.00001 |
| BMI | 21.81±1.94 | 21.13±4.74 | n.s. |
| FBG (mg/dl) | 228.1±62.5 | 191.0±64.1 | <0.008 |
| HbA1c (%) | 10.89±2.61 | 9.22±2.94 | <0.0002 |
| insulin (μU/ml) | 4.91±2.82 | 5.25±3.07 | n.s. |
| FFA (mEq/L) | 0.776±0.399 | 0.726±0.334 | n.s. |
| HOMA-R | 2.91±2.13 | 2.59±2.16 | n.s. |
| adipo-IR | 5.69±2.98 | 3.86±3.12 | <0.02 |
| HOMA-B | 11.64±6.12 | 17.80±11.39 | <0.006 |

BMI≥25: group Y
|  | baseline | 3 months | p values |
| --- | --- | --- | --- |
| N | 26 |  |  |
| age | 48.9±12.2 |  |  |
| UA (mg/dl) | 5.35±1.52 | 5.85±1.61 | <0.02 |
| BMI | 28.44±3.84 | 28.41±3.62 | n.s. |
| FBG (mg/dl) | 227.4±60.4 | 192.6±63.2 | <0.01 |
| HbA1c (%) | 10.50±1.84 | 8.80±1.77 | <0.0002 |
| insulin (μU/ml) | 8.13±5.64 | 9.38±5.35 | n.s. |
| FFA (mEq/L) | 0.833±0.309 | 0.747±0.262 | n.s. |
| HOMA-R | 4.72±3.71 | 4.65±3.50 | n.s. |
| adipo-IR | 8.96±5.62 | 7.19±5.60 | n.s. |
| HOMA-B | 19.09±11.95 | 30.43±18.31 | <0.004 |

**Figure 1.**
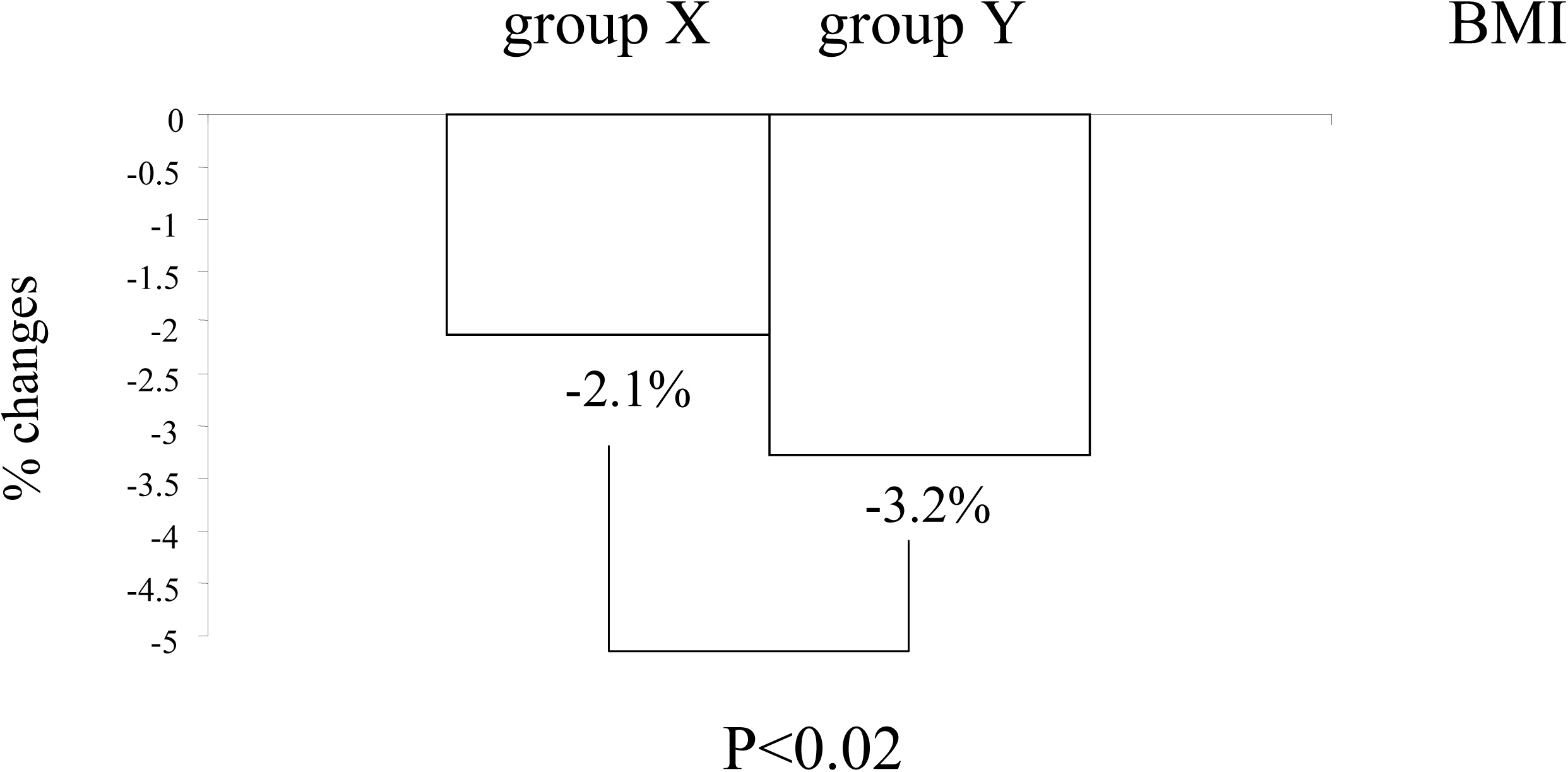
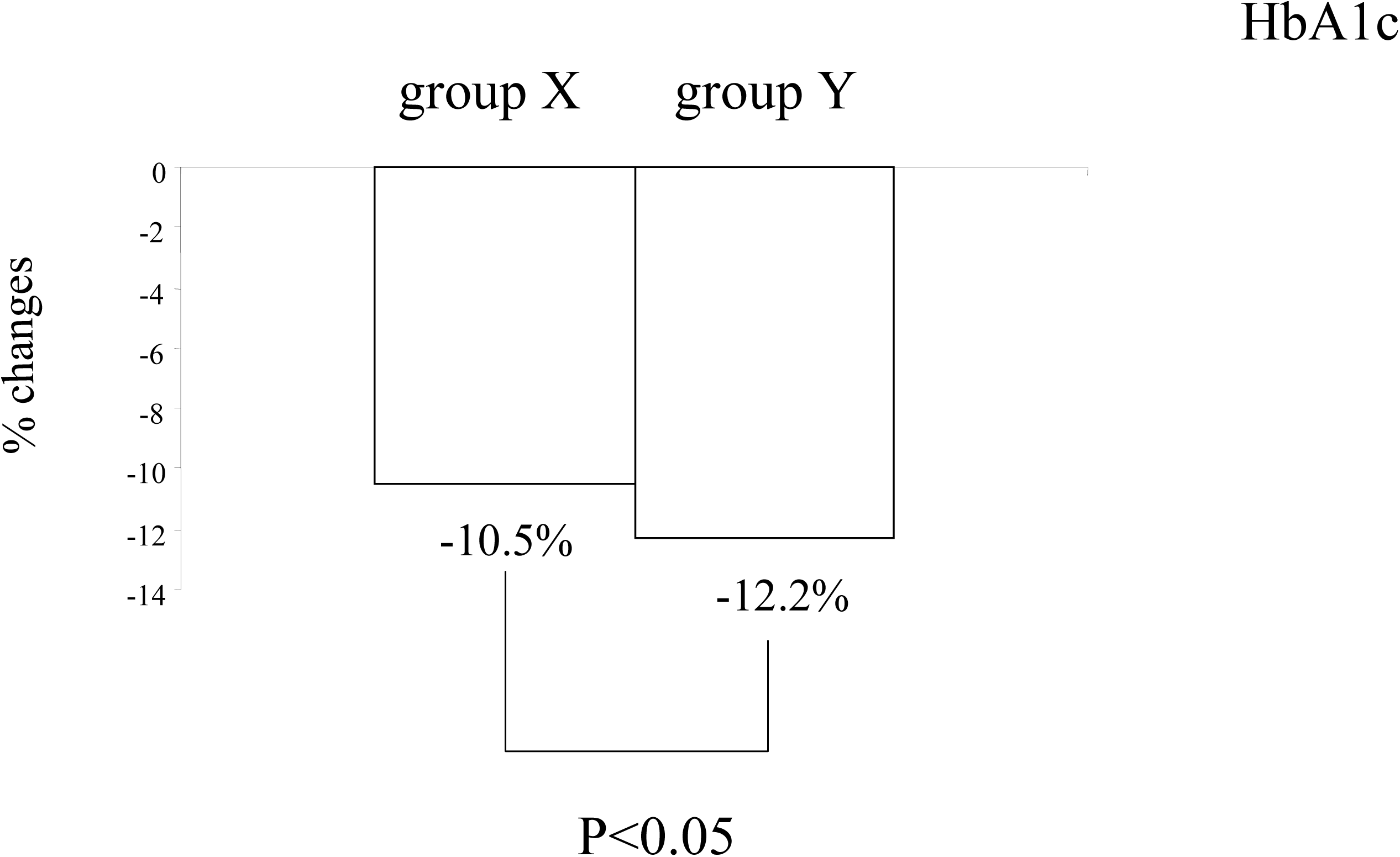
Differential effects on diabetic parameters by the baseline weight (BMI<25 and BMI>25) ANCOVA was performed to analyze the inter-group differences on the changes of the indicated parameters in the MJDD group (% changes). Panel A) BMI Panel B) HbA1c

By contrast with alogliptin, in group X (Table 3B, n=26), significant decreases of FBG, HbA1c or adipo-IR, and increases of UA and HOMA-B were seen. In group Y (Table 3B, n=26), significant decreases of FBG or HbA1c, and increases of UA and HOMA-B were observed. No inter-group differences were noted with UA and HOMA-B in these two groups (X and Y, results not shown as Figure).

### Correlations Between UA and Diabetic Parameters at Baseline and After 3 Months

At baseline, significant correlations were found between UA and BMI, insulin, HOMA-R, adipo-IR and HOMA-B in both groups (Table 4A: MJDD and 4B: alogliptin). By contrast, no significant correlations were observed between UA and FBG, HbA1c or FFA (Table 4A, 4B). At 3 months, significant positive correlations between ΔUA and ΔHOMA-B, and negative correlations between ΔUA and ΔHbA1c were observed in both groups (Table 5A, 5B). Additional positive correlations between ΔUA and ΔFFA, and negative correlations between ΔUA and ΔFBG or ΔBMI were seen only in MJDD group (Table 5A).

**Table 4.**
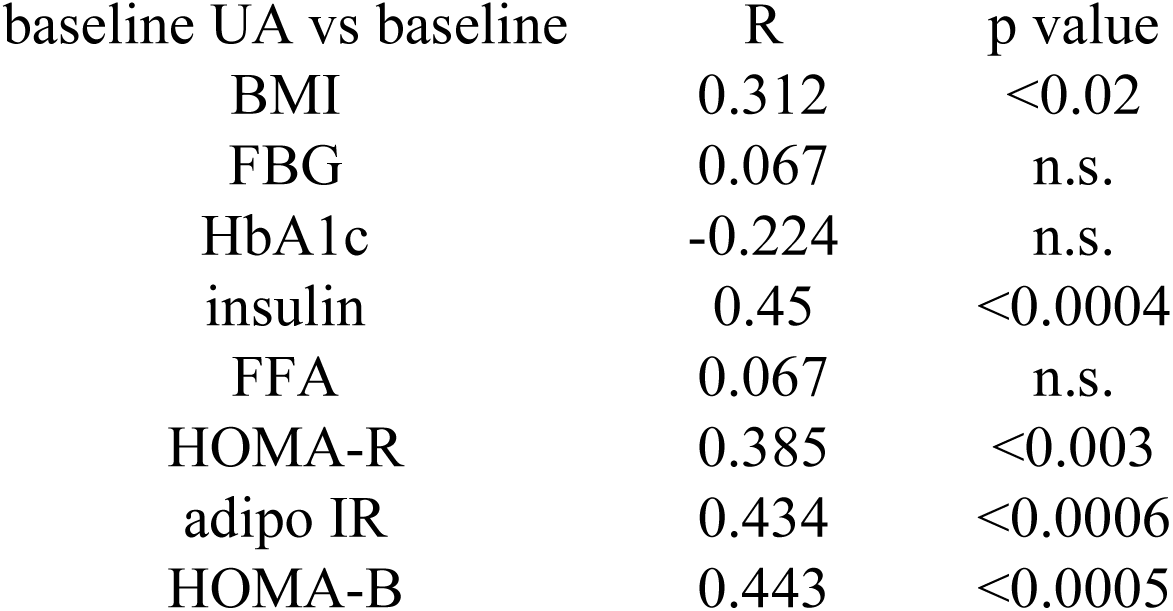
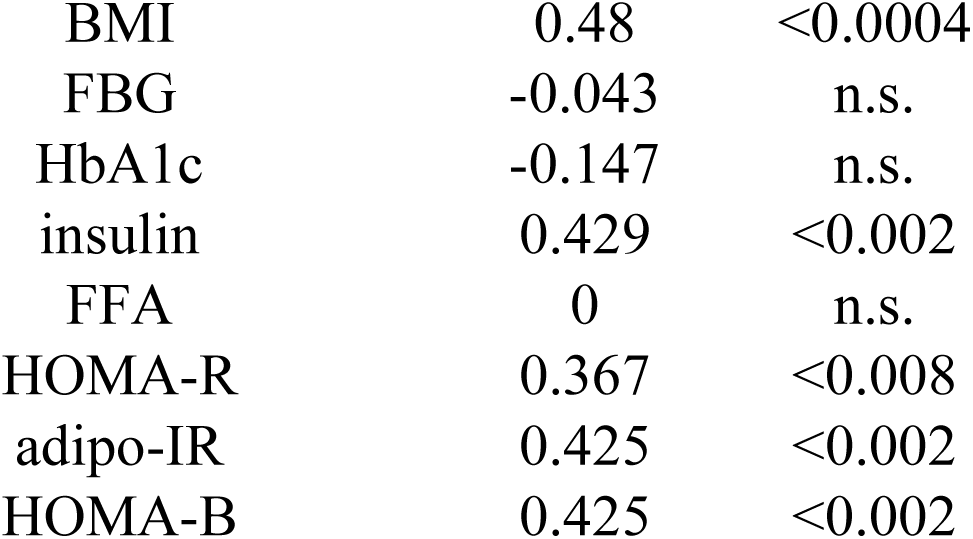
Correlation between the baseline levels of UA and those of other diabetic parameters with MJDD and alogliptin. Simple regression analysis was performed between the baseline levels of UA and those of diabetic parameters. A) MJDD B) alogliptin

| baseline UA vs baseline | R | p value |
| --- | --- | --- |
| BMI | 0.312 | <0.02 |
| FBG | 0.067 | n.s. |
| HbA1c | -0.224 | n.s. |
| insulin | 0.45 | <0.0004 |
| FFA | 0.067 | n.s. |
| HOMA-R | 0.385 | <0.003 |
| adipo IR | 0.434 | <0.0006 |
| HOMA-B | 0.443 | <0.0005 |
| BMI | 0.48 | <0.0004 |
| FBG | -0.043 | n.s. |
| HbA1c | -0.147 | n.s. |
| insulin | 0.429 | <0.002 |
| FFA | 0 | n.s. |
| HOMA-R | 0.367 | <0.008 |
| adipo-IR | 0.425 | <0.002 |
| HOMA-B | 0.425 | <0.002 |

**Table 5.**
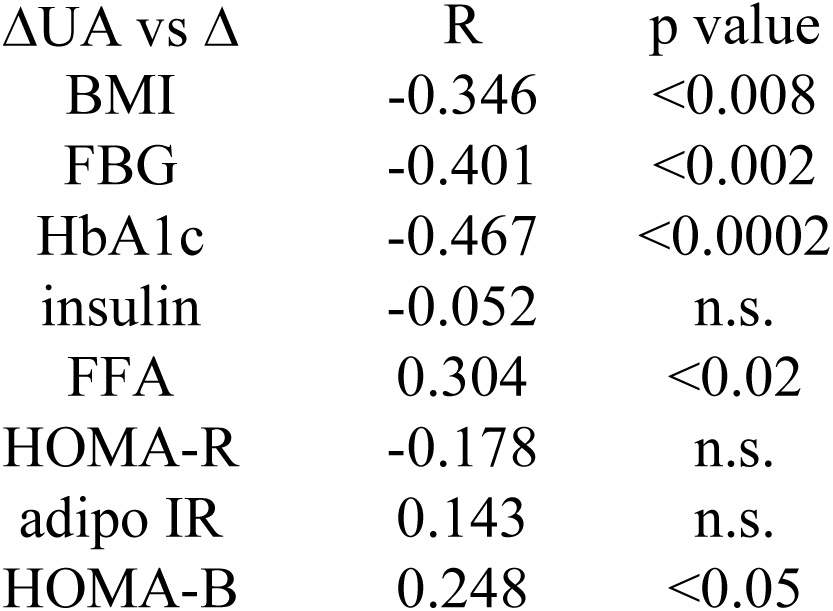
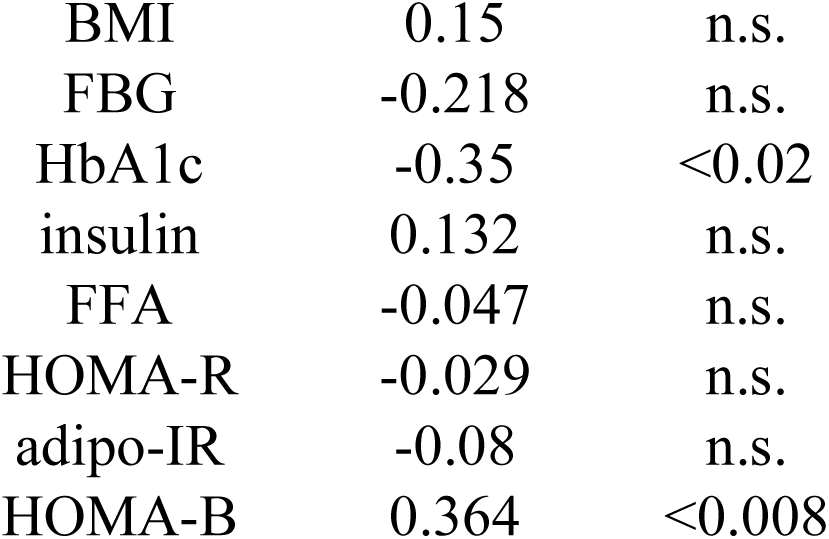
Correlation between the changes of UA and those of other diabetic parameters with MJDD and alogliptin. A) MJDD B) alogliptin Simple regression analysis was performed between the changes of (Δ) UA and those of diabetic parameters.

| ΔUA vs $\Delta$ | R | p value |
| --- | --- | --- |
| BMI | -0.346 | <0.008 |
| FBG | -0.401 | <0.002 |
| HbA1c | -0.467 | <0.0002 |
| insulin | -0.052 | n.s. |
| FFA | 0.304 | <0.02 |
| HOMA-R | -0.178 | n.s. |
| adipo IR | 0.143 | n.s. |
| HOMA-B | 0.248 | <0.05 |
| BMI | 0.15 | n.s. |
| FBG | -0.218 | n.s. |
| HbA1c | -0.35 | <0.02 |
| insulin | 0.132 | n.s. |
| FFA | -0.047 | n.s. |
| HOMA-R | -0.029 | n.s. |
| adipo-IR | -0.08 | n.s. |
| HOMA-B | 0.364 | <0.008 |

### Differential Regulation of Diabetic Parameters Based on UA Changes

Subjects in each treatment group were stratified into two subgroups according to the median value of UA change (ΔUA): group X (lower half) and group Y (upper half), as described in the Subjects and Methods.

#### 1) MJDD

Group X (N=33): Significant reductions in UA, BMI, HbA1c, FFA or adipo-IR and insignificant reductions of insulin were observed (Table 6A).

**Table 6.** Regulations of diabetic parameters depending on the changes of UA with MJDD and alogliptin. The subjects were divided into two groups based on the median changes of (Δ)UA (lower half: group X and upper half: group Y) in these two therapeutic groups. Paired Student’s t-test was used to compare the changes in the indicated parameters at baseline and 3 months treatment. The results are expressed as the mean±standard deviation (SD) A) MJDD B) alogliptin

group X
|  | baseline | 3 months | p values |
| --- | --- | --- | --- |
| N | 33 |  |  |
| age | 52.3±11.7 |  |  |
| UA (mg/dl) | 5.13±1.37 | 4.96±1.31 | <0.005 |
| BMI | 25.05±4.73 | 24.55±4.69 | <0.004 |
| FBG (mg/dl) | 177.9±45.6 | 169.4±48.4 | n.s. |
| HbA1c (%) | 8.71±1.50 | 8.03±1.54 | <0.0008 |
| insulin (μU/ml) | 7.00±4.64 | 6.06±3.23 | 0.078 |
| FFA (mEq/L) | 0.808±0.436 | 0.610±0.221 | <0.008 |
| HOMA-R | 3.20±2.55 | 2.56±1.68 | <0.04 |
| adipo-IR | 5.40±4.13 | 3.88±3.17 | <0.004 |
| HOMA-B | 23.66±14.69 | 24.75±15.89 | n.s. |

group Y
|  | baseline | 3 months | p values |
| --- | --- | --- | --- |
| N | 25 |  |  |
| age | 48.1±12.6 |  |  |
| UA (mg/dl) | 5.01±1.42 | 5.74±1.42 | <0.00001 |
| BMI | 26.49±4.61 | 25.55±4.41 | <0.00001 |
| FBG (mg/dl) | 175.0±53.1 | 154.5±57.0 | <0.002 |
| HbA1c (%) | 8.89±1.32 | 7.44±1.33 | <0.00001 |
| insulin (μU/ml) | 8.66±5.41 | 8.06±4.67 | n.s. |
| FFA (mEq/L) | 0.674±0.223 | 0.700±0.244 | n.s. |
| HOMA-R | 3.65±2.33 | 2.93±1.75 | <0.02 |
| adipo-IR | 5.79±3.90 | 5.81±4.24 | n.s. |
| HOMA-B | 33.32±23.18 | 41.71±29.99 | <0.02 |

| group X |  |  |  |
| --- | --- | --- | --- |
|  | baseline | 3 months | p values |
| N | 25 |  |  |
| age | 53.9±13.8 |  |  |
| UA (mg/dl) | 4.88±1.79 | 4.73±1.61 | n.s. |
| BMI | 25.65±4.32 | 24.41±6.49 | n.s. |
| FBG (mg/dl) | 219.5±66.8 | 201.5±63.3 | <0.05 |
| HbA1c (%) | 10.45±2.37 | 9.56±2.54 | <0.007 |
| insulin (μU/ml) | 6.33±4.64 | 6.36±4.21 | n.s. |
| FFA (mEq/L) | 0.795±0.322 | 0.777±0.309 | n.s. |
| HOMA-R | 3.54±3.18 | 3.27±2.70 | n.s. |
| adipo-IR | 7.12±4.67 | 5.11±4.67 | n.s. |
| HOMA-B | 16.32±10.15 | 19.06±11.75 | n.s. |

| group Y |  |  |  |
| --- | --- | --- | --- |
|  | baseline | 3 months | p values |
| N | 27 |  |  |
| age | 51.8±12.6 |  |  |
| UA (mg/dl) | 4.47±1.30 | 5.64±1.48 | <0.00001 |
| BMI | 24.67±4.69 | 25.07±4.73 | 0.054 |
| FBG (mg/dl) | 234.8±55.5 | 183.5±62.7 | <0.002 |
| HbA1c (%) | 10.91±2.16 | 8.55±2.24 | <0.00001 |
| insulin (μU/ml) | 6.68±4.84 | 8.14±5.18 | n.s. |
| FFA (mEq/L) | 0.813±0.386 | 0.702±0.287 | n.s. |
| HOMA-R | 4.05±3.12 | 3.93±3.35 | n.s. |
| adipo-IR | 7.50±4.90 | 5.89±4.95 | n.s. |
| HOMA-B | 14.54±10.22 | 28.44±18.65 | <0.0003 |

Group Y (N=25): Significant reductions in BMI, FBG, HbA1c, or HOMA-R and increases in UA and HOMA-B were observed (Table 6B). Significant inter-group differences in the changes in BMI and HbA1c were seen, with greater reductions in group Y versus group X (Figure 2A, 2B).

**Figure 2.**
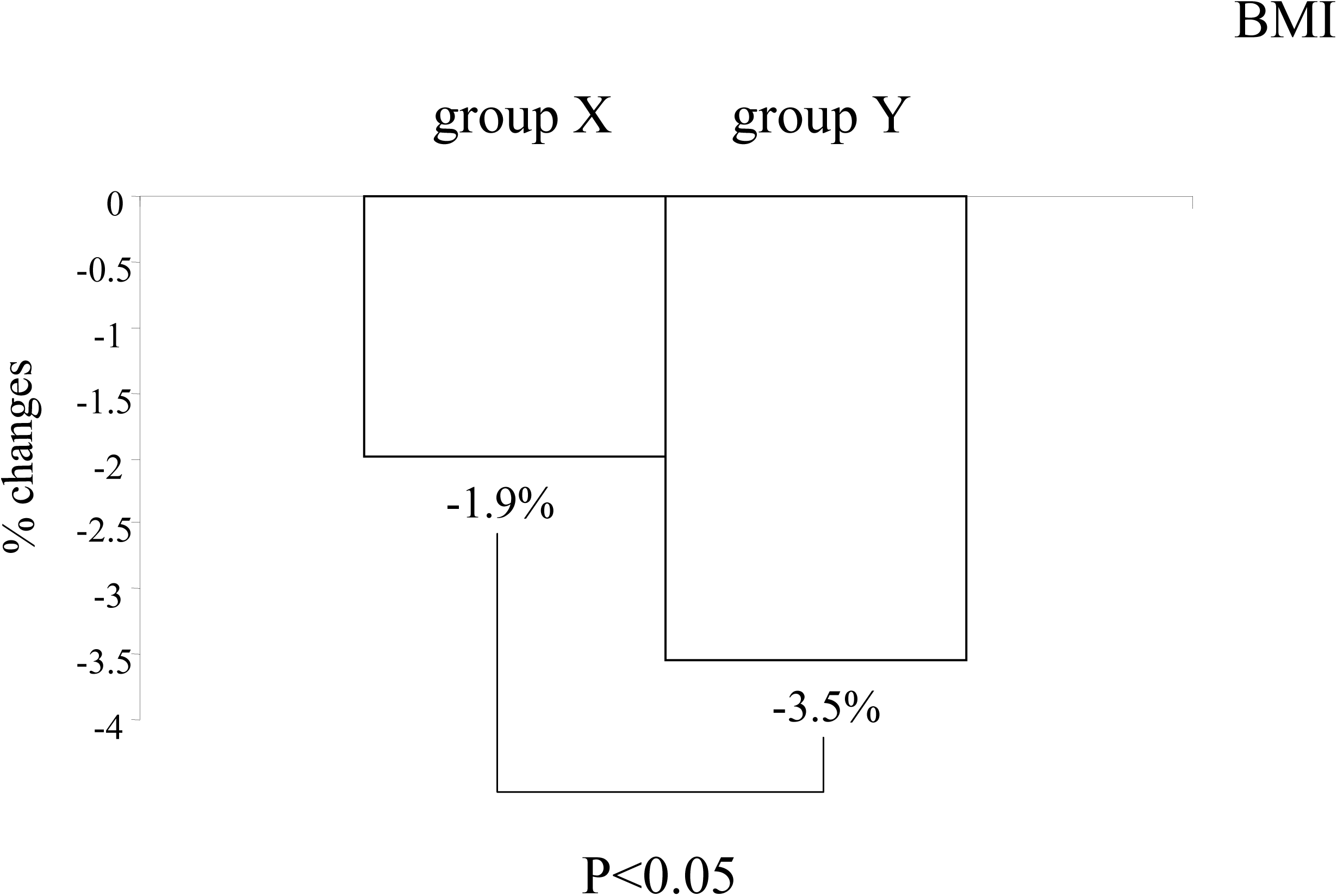
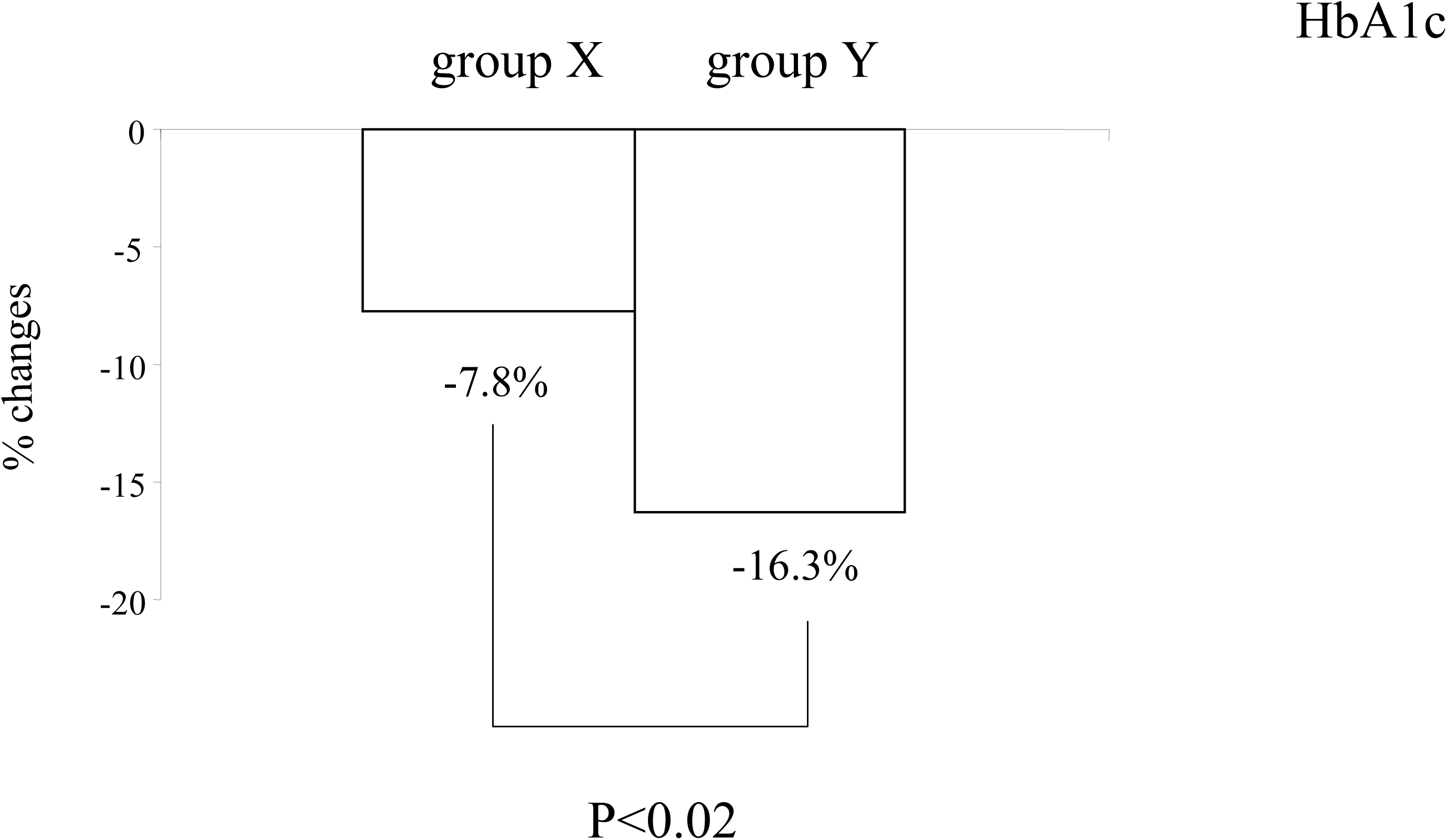
Differential effects on diabetic parameters by the median changes of (Δ) UA (group X: lower half group Y: upper half MJDD) ANCOVA was performed to analyze the inter-group differences on the changes of the indicated parameters in the MJDD group (% changes). Panel A) BMI Panel B) HbA1c

#### 2) Alogliptin

Group X (N=25): Significant reductions in FBG and HbA1c were observed (Table 6B).

Group Y (N=27): Significant reductions in FBG and HbA1c, along with increases in UA and HOMA-B, were observed. BMI had a tendency to increase (Table 6B). Significant inter-group differences in the changes in FBG and HbA1c were seen with greater reductions in group Y versus group X (Figure 3A, 3B).

**Figure 3.**
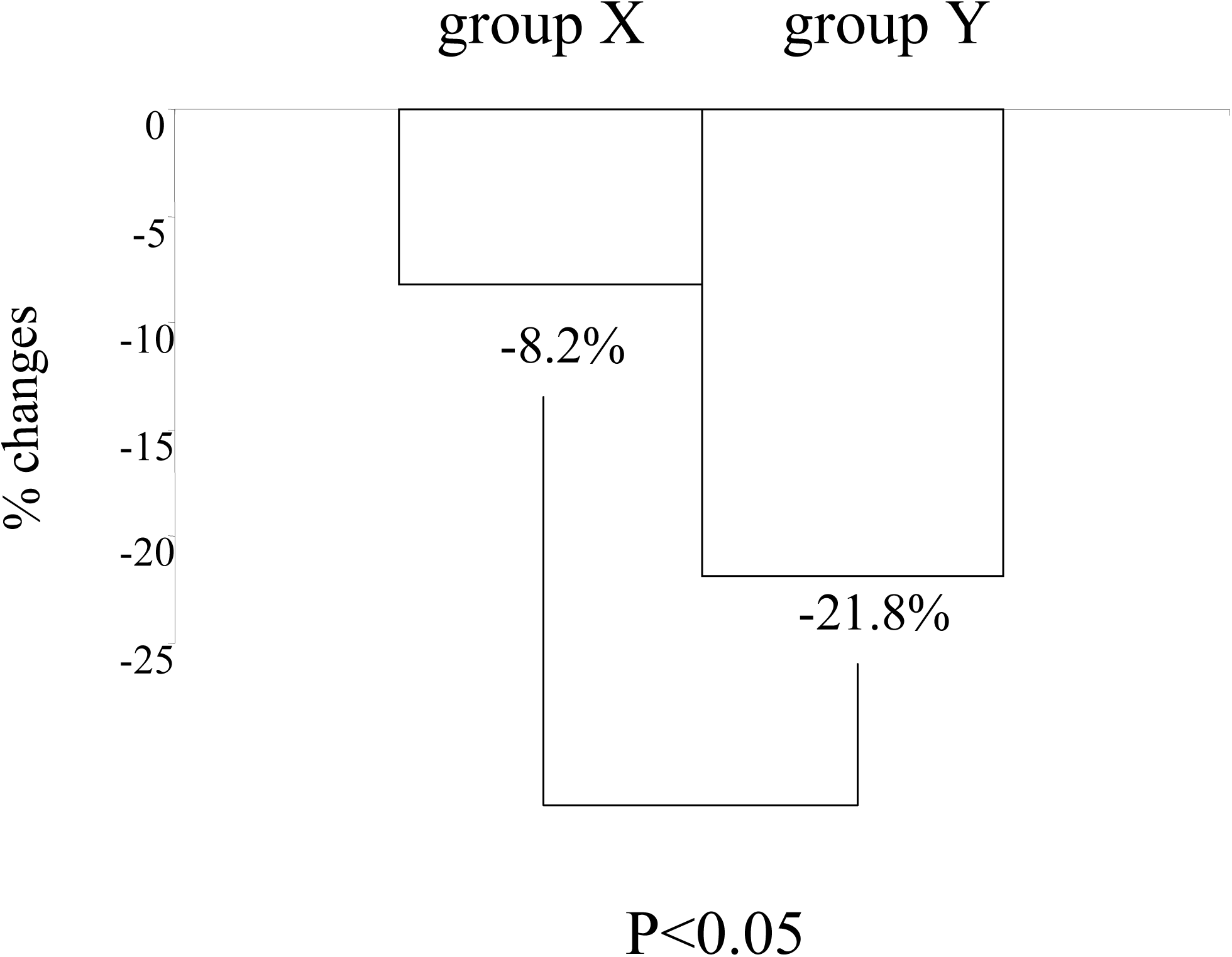
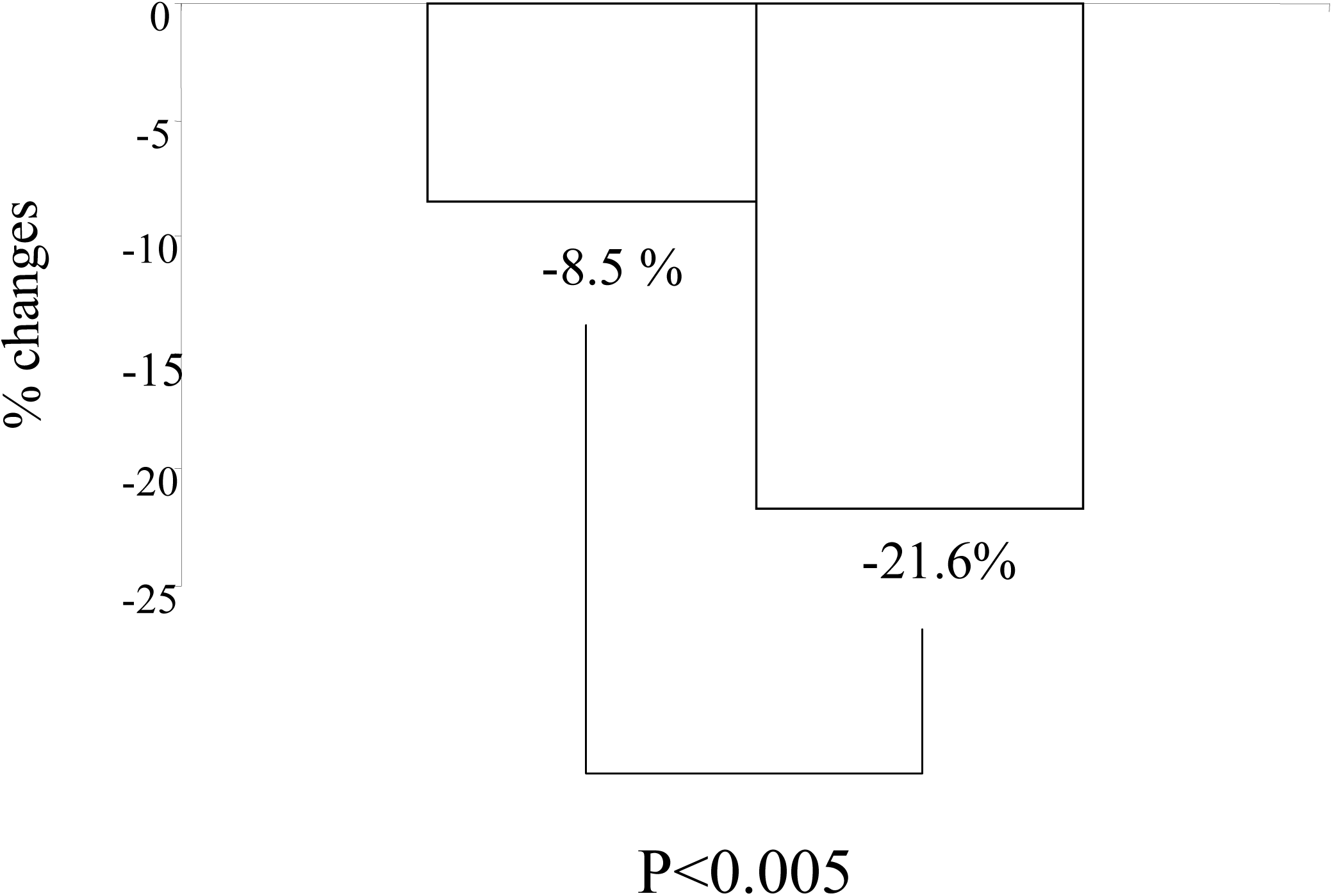
Differential effects on diabetic parameters by the median changes of (Δ) UA (group X: lower half, group Y: upper half alogliptin) ANCOVA was performed to analyze the inter-group differences on the changes of the indicated parameters in the alogliptin group (% changes). Panel A) FBG Panel B) HbA1c

## Discussion

### Weight dependent differential regulation of diabetic parameters during treatment with modified traditional Japanese diet for diabetes (MJDD) and DPP-4 inhibitor alogliptin depending on baseline weight

This study provides novel insights into how UA dynamics reflect distinct metabolic adaptations under two therapeutic strategies: MJDD and the DPP-4 inhibitor alogliptin. Our primary finding was that both interventions improved glycemic control and enhanced beta-cell function while simultaneously elevating serum UA levels (Table 1A and 1B). This is particularly striking given the divergent ways these therapies have distinct impact on body weight and insulin resistance (Table 3A, 3B, Fig. 1, 2). While alogliptin is primarily recognized for its glucose-dependent insulinotropic effects, our findings suggest it also modulates uric acid (UA) dynamics. The observed increase in UA following alogliptin therapy, which correlated significantly with ΔHOMA-B, suggests that DPP-4 inhibition may influence purine metabolism or renal UA handling secondary to improvements in beta-cell secretory demand. This underscores a potential pleiotropic effect of alogliptin that warrants further investigation into transporter-level interactions. In contrast, the UA elevation seen with MJDD occurred despite significant weight loss, suggesting a different, perhaps more complex, metabolic decoupling.

Baseline UA appears to be the determinant of the changes of UA in these therapeutic strategies (Table 2A, 2B). With MJDD, the subjects with higher BMI (≥25, group Y) experienced pronounced reductions in glycemic parameters, (adipose tissue) insulin resistance, FFA, and body weight, with minimal changes in UA and beta-cell function (Table 3A, Fig.1A, 1B). Conversely, the subjects with lower BMI (<25, group X) exhibited notable increases in UA and beta-cell function but showed minimal improvements in insulin resistance or glycemic markers (Table 3A, Fig. 1B). This discordance suggests that in higher-weight individuals, the glycemic efficacy of this dietary therapy is predominantly mediated via insulin sensitization, whereas in lower weight individuals, beta-cell enhancement predominates despite lesser metabolic and glycemic benefit. By contrast, alogliptin improved glycemic parameters primarily through augmentation of beta-cell function, consistent with GLP-1-mediated effects (12, 13), with minimal impact on weight or whole body insulin sensitivity (Table 3B).

One interesting, novel observation in this study is the differential regulation of adipose tissue insulin resistance (adipo-IR) depending on the baseline weight (Table 3A, 3B). This BMI-stratified analysis indicates that MJDD and alogliptin improve adipose tissue insulin resistance (adipo-IR) through distinct, BMI-dependent mechanisms. With MJDD, a significant reduction in adipo-IR was observed only in subjects with higher BMI (≥25), consistent with effects targeting obesity-related adipose dysfunction (group Y, Table 3A). In subjects with lower BMI (<25), baseline adipose dysfunction is likely milder, leaving less scope for measurable improvement in adipo-IR (group X, Table 3A); in these with lower BMI subjects, the dietary intervention may exert its main benefits on glycemic control and beta-cell function rather than adipose tissue insulin sensitivity. By contrast, alogliptin reduced adipo-IR only in the lower BMI group (group X, Table 3B), suggesting a different pathway dominated by hormonal modulation. DPP-4 inhibition enhances incretin action, particularly GLP-1 and GIP [13], which may be more effective in improving adipose tissue insulin sensitivity when adipose tissue remains relatively free of structural alterations such as fibrosis or severe hypertrophy. In those with lower BMI, the GLP-1 and other hormonal pathways may remain intact and responsive, whereas in individuals with higher BMI, chronic inflammation and structural remodeling of adipose tissue may blunt these effects. This weight-dependent divergence supports the concept of therapeutic phenotype matching, favoring diet-first strategies for obese patients with marked adipose insulin resistance, and incretin-based therapy for leaner patients in whom adipose tissue dysfunction is present but primarily functional rather than structural.

### Link between UA and diabetic parameters during treatment with modified traditional Japanese diet (MJDD) and DPP-4 inhibitor alogliptin

Baseline correlations between UA and some diabetic parameters indicate that UA primarily reflects underlying insulin resistance, weight and beta-cell demand, rather than short-term glycemic or lipid (FFA) status (Table 4A, 4B). Correlation between the changes of (Δ)UA and those of diabetic parameters and stratification by the changes of (Δ)UA with MJDD group revealed that increased UA was associated with improved beta-cell function and glycemic markers, whereas decreased UA paralleled improvements in adipose tissue insulin resistance (adipo-IR) or FFA, but not in whole body insulin resistance (HOMA-R) or weight (Table 5A, 6A, irrespective of the changes of UA, significant reductions of BMI or HOMA-R were seen in these two subgroups). These findings indicate a potential link between UA and adipose tissue metabolism and at the same time decoupling between UA and whole body insulin resistance or weight under certain conditions, such as during treatment with MJDD. This could be a novel mode of UA regulation during metabolic improvements.

With alogliptin, correlation between the changes of (Δ)UA and those of diabetic parameters and stratification by the changes of UA within this drug group revealed that UA changes showed that increased UA was associated with enhanced beta-cell function and greater reductions in glycemic indices (Table 5B, 6B).

Taken together, these observations suggest that UA may serve as a surrogate biomarker reflecting the dominant metabolic pathway activated by a given therapy-insulin sensitization vs. beta-cell activation-rather than simply mirroring purine metabolism or renal handling (14). The contrasting effects of diet and pharmacologic therapy on adipo-IR and UA further support this interpretation and underscore the relevance of precision medicine in early T2DM management.

### Potential mechanism of paradoxical increase of UA with modified traditional Japanese diet for diabetes (MJDD)

Interestingly, despite traditional Japanese diet inducing significant reductions in body weight, insulin resistance (HOMA-R), and FFA-all of which are typically associated with improved UA handling (15)-UA levels paradoxically increased (Table 1A).

There are a number of explanations for these unexpected results:

1. Traditional Japanese diet contains purine-rich components such as fish, seafood, and their organs. This is supported by HPLC-based purine analyses of 270 Japanese food items (16).
2. Weight loss, particularly through reduction in adipose tissue mass, may enhance purine degradation, thereby increasing UA production. Such mechanisms may be particularly relevant during the initial stages of dietary intervention before metabolic equilibrium is re-established. This partially, if not fully, explains the unexpected rise in UA with concomitant weight loss and the negative correlation between ΔUA and ΔBMI. To support this idea, with MJDD, higher degrees of weight loss were observed in those of increases of UA in comparison to others of decreased UA (group Y versus X, Fig. 2A).
3. Since glycosuria promotes uric acid excretion by competing with UA for reabsorption via transporters such as GLUT9 and URAT1 (17), reduced glycosuria by improved blood glucose levels with this therapy may decrease UA clearance. Thus, the uricosuric effect of hyperglycemia is lost, leading to a net increase in serum UA despite favorable changes in metabolic parameters. Significant increases of UA with DPP-4 inhibitor alogliptin (Table 1B) may be partially, not fully, due to this mechanism as well.
4. Significant reductions of FFA were observed with MJDD therapy (Table 1A). Since FFA can compete with UA for renal excretion via shared transporters such as URAT1 and GLUT9 (18), a reduction in circulating FFA levels may decrease this competition, thereby impairing UA excretion and increasing serum UA concentrations. Consequently, individuals who experienced greater increases in UA showed greater weight loss and improvement in adipose tissue insulin resistance (with concurrent FFA reduction), leading to the observed negative correlations (Table compare group X versus group Y, Table 6A, Fig. 2A). Emerging evidence highlights that metabolic intermediates such as FFA, lactate, and ketone bodies, all of which are elevated during catabolic states or acute weight loss, can competitively inhibit UA secretion through OAT1 and OAT3 transporters (19). Such substrate competition could contribute to reduced UA clearance and underlie the negative correlations observed between changes in UA and changes in BMI with MJDD (Table 5A).

Notably, while baseline levels of UA and FFA were uncorrelated, significant correlations between ΔUA and ΔFFA with the treatment of MJDD (Table 4A, 5A) suggest that this therapy induced a coordinated metabolic response. Suppression of lipolysis (reductions of FFA in other words) by improved adipose tissue insulin sensitivity together with enhanced fatty acid oxidation may have increased purine turnover and UA production. The observed association between ΔFFA and ΔUA likely reflects this shift toward lipid-based metabolism and nucleotide degradation under caloric restriction, rather than changes in renal UA excretion. These findings highlight a dynamic, intervention-dependent link between FFA and UA metabolism that is not evident at baseline.

### Enhancement of beta-cell function with UA

In both therapeutic strategies, beta-cell function (assessed using HOMA-B) significantly improved, and changes in (Δ)HOMA-B were positively correlated with ΔUA. Moreover, ΔHbA1c were negatively correlated with ΔUA (Table 1A, 1B, 5A, 5B). These results suggest that elevations in UA may accompany both enhanced insulin secretory capacity and improved glycemic control. Stratified analyses further revealed that subjects with greater increases in UA (Group Y in each treatment arm; Tables 6A, 6B) demonstrated more pronounced improvements in beta-cell function and achieved better glycemic outcomes, as illustrated in Fig. 2B, 3A, 3B). These findings strongly support a physiological link between UA and beta-cell function. While the mechanisms by which UA may enhance beta-cell function remain largely unknown, several hypotheses can be considered. One possibility is that UA, particularly in moderate concentrations, acts as a physiological antioxidant, counteracting oxidative stress within pancreatic islets (20). Since beta-cells are particularly susceptible to oxidative damage due to their relatively low expression of antioxidant enzymes (21, 22), the protective effect of UA may help preserve or even enhance insulin secretory capacity. In addition, UA may influence metabolic or inflammatory pathways-such as the activation of Nrf2 signaling or suppression of pro-inflammatory cytokines (e.g., IL-1β, TNF-α)-that are known to affect beta-cell survival and function (23, 24, 25). It is also plausible that elevated UA reflects a state of increased nucleotide turnover associated with greater energy availability, which might indirectly support beta-cell metabolic activity and insulin synthesis. However, whether UA itself exerts direct protective effects on beta-cells or is simply a marker of other underlying metabolic changes remains to be elucidated. Future mechanistic and interventional studies are needed to clarify the causality and therapeutic relevance of this relationship.

### Limitations and future perspectives of this study

This study has several limitations. It is an observational study with a relatively small sample size and a short study duration. While randomized controlled trials (RCTs) are considered the gold standard for evaluating the efficacy of interventions due to their ability to minimize bias and confounding, However, observational studies have inherent advantages, including reflecting real-world conditions and potentially capturing natural randomization effects. Moreover, our study protocol, which involved monotherapy in drug-naïve subjects, strengthens the attribution of observed effects to the interventions employed. Further, we did not measure urinary uric acid excretion or renal function parameters in detail, which limits mechanistic interpretation regarding uric acid handling. Lastly, the lack of blinding and potential selection bias should be considered when interpreting subgroup differences.

## Conclusion

Collectively, these findings support the hypothesis that dietary intervention, particularly in leaner individuals, may elevate UA levels in parallel with improved beta-cell function and glycemic control, whereas heavier individuals may experience UA stabilization due to concurrent improvements in insulin resistance and weight loss. The differential patterns between diet and drug interventions underscore the complex and context-dependent regulation of UA metabolism in T2DM. We hypothesize that the effects of anti-diabetic therapies on UA metabolism are modulated by distinct regulatory pathways that go beyond weight reduction and insulin sensitivity. UA emerged as a dynamic marker reflecting these divergent responses. These findings support the value of incorporating metabolic profiling—such as baseline BMI and uric acid levels—into individualized therapeutic strategies for T2DM.

## Data Availability

All data produced in the present study are available upon reasonable request to the authors.

## Conflicts of Interest

The authors declare that no conflicts of interest exist regarding this manuscript.

CRediT authorship contribution statement: E.K. and A.N.K. participated in the design of the study and acquisition of the data, performed the statistical analysis, and drafted the manuscript. N.U, R.O., and S.I. made substantial contributions to the conception and design of the study and helped draft the manuscript. Writing, review and editing: E.K. and A.N.K. All authors have read and agreed to the published version of the manuscript.

## Funding

This research received no external funding.

## Informed Consent Statement

Informed consent was obtained from all patients and stored in the electronic medical record system.

## Data Availability Statement

The data that support the findings of this study are available from the corresponding author (E.K.) upon reasonable request.

## Acknowledgments

The authors express gratitude to Asuka Wada, Midori Akiyama and Rumi Kurihara for their insightful discussions and valuable feedbacks. The authors also appreciate Keiko Saido-Ozawa for her administrative support.

## Abbreviations

T2DM: type 2 diabetes
MJDD: modified Japanese diet for diabetes
DPP-4: dipeptidyl peptidase 4
BMI: body mass index
adipo-IR: adipose tissue insulin resistance
FBG: fasting blood glucose
HOMA-R: homeostasis model assessment-R
HOMA-B: homeostasis model assessment-B

## Notes

### Competing Interest Statement

The authors have declared no competing interest.

### Clinical Trial

UMIN000006860

### Funding Statement

This study did not receive any funding.

### Author Declarations

The study was conducted in accordance with the Declaration of Helsinki and was approved by the institutional review board (IRB) of Gyoda General Hospital and Kumagaya Surgery Hospital. Written informed consent was obtained and recorded electronically. This project is an extension of the registered study (ID: UMIN000006860), which is listed in the UMIN database.

